# Impact of the accuracy of case-based surveillance data on the estimation of time-varying reproduction numbers

**DOI:** 10.1101/2020.06.26.20140871

**Authors:** Michele Starnini, Alberto Aleta, Michele Tizzoni, Yamir Moreno

**Affiliations:** ISI Foundation, via Chisola 5, 10126, Torino, Italy; Institute for Biocomputation and Physics of Complex Systems (BIFI), University of Zaragoza, Spain; Department of Theoretical Physics, Faculty of Sciences, University of Zaragoza, Spain

**Keywords:** COVID-19, time-varying reproductive number, public health

## Abstract

Studies aimed at characterizing the evolution of COVID-19 disease often rely on case-based surveillance data publicly released by health authorities, that can be incomplete and prone to errors. Here, we quantify the biases caused by the use of inaccurate data in the estimation of the Time-Varying Reproduction Number R(t). By focusing on Italy and Spain, two of the hardest-hit countries in Europe and worldwide, we show that if the symptoms’ onset time-series is inferred from the notification date series, the R(t) curve cannot capture nor describe accurately the early dynamics of the epidemic. Furthermore, the effectiveness of the containment measures that were implemented, such as national lockdowns, can be properly evaluated only when R(t) is estimated using the real time-series of dates of symptoms’ onset. Our findings show that extreme care should be taken when a pivotal quantity like R(t) is used to make decisions and to evaluate different alternatives.

## Introduction

The COVID-19 pandemic has brought the time-varying reproduction number, R(t), to the public attention worldwide [1]. R(t) represents the mean number of secondary infections generated by one primary infected individual, over the course of an epidemic. In contrast to the basic reproductive number, R0 [2], which assumes the population to be fully susceptible, the temporal variations of R(t) reflect the changes in transmissibility caused by the depletion of susceptibles during the epidemic, and by the presence of control interventions and spontaneous behavioral responses. When R(t) decreases below the threshold value of 1, the number of new infections begins to decline. This quantity is thus pivotal to evaluate the impact of interventions, calibrate epidemiological models, and guide the lift of restrictions, where in place.

Usually, R(t) can be estimated by means of various inference algorithms applied to epidemic incidence data (the numbers of new recorded cases at successive times). Although a precise estimate of R(t) would require knowledge of the onset of symptoms for all cases belonging to the incidence curve, many published estimates of R(t) during the COVID-19 pandemic have relied on incidence curves reporting cases by notification date, since the true date of symptoms’ onset is usually unknown or it is not publicly available [3,4]. Several estimation methods account for such lack of information by using time-independent delay distributions from symptom onset to reporting [5,6]. However, the impact of using case-notification data - instead of the true symptom onset - as the main data input for estimating R(t) has not been investigated so far.

## Data

To address this issue, we evaluate R(t) by using four different types of incidence curves inferred from different data sets. We focus on two European countries that are among the most severely hit by the COVID-19 pandemic, Italy and Spain. First, we consider the time-series of laboratory-confirmed COVID-19 cases by the date of symptoms’ onset, from early February to late May as reported by national health authorities. In Italy, this data is available solely in graphic format in the reports published by the Istituto Superiore di Sanità

(ISS), whereas Spain has made this data publicly available only recently in machine-readable format. Secondly, we use the time-series of reported cases by notification date, as provided by the European Centre for Disease Prevention and Control (ECDC). The third time-series of symptoms’ onset is inferred (Inferred Symptoms A) by applying a convolution to the time-series of notification dates with the probability distribution P(Δt) of the elapsed time between the dates of symptom onset and laboratory confirmation, measured from a large line list dataset [7]. Lastly, another time-series of symptoms’ onset could be inferred (Inferred Symptoms B) by backwards shifting the dates of laboratory-confirmed deaths by 11 days, which corresponds to the median time elapsed between symptom onset and death in Spain^1^ and Italy^2^. The resulting time-series of real symptoms’ onset, notification date and inferred symptoms’ onset A and B are shown in Fig. 1.

**Figure 1.**
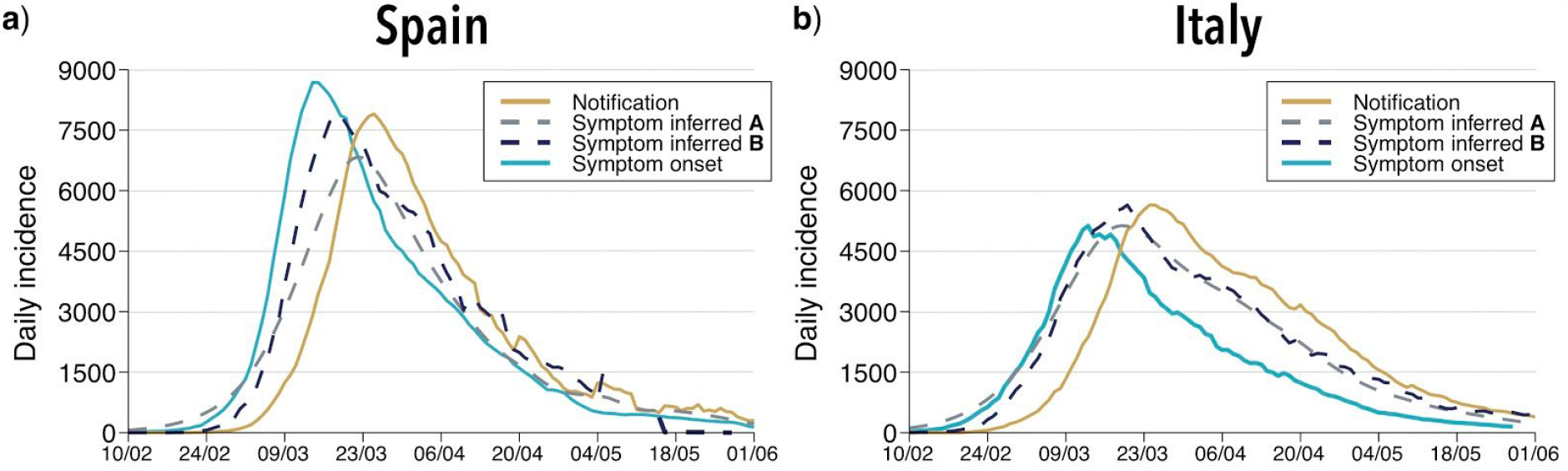
Time-series of the incidence by real symptom onset, notification date, inferred symptom onset by convolution with P(Δt) and time-shifted confirmed deaths in Spain (left) and Italy (right). Time-series are smoothed with a moving average of 7 days.

One can see how the time-series of the time-shifted deaths, while obtained with a rough approximation, is remarkably similar to the time-series of the symptom onset dates inferred through the delay distribution P(Δt). However, both curves fail to match the true epidemic curve by symptom onset, which peaks earlier and declines faster than the two inferred curves. Note also that there is a significant delay between the time-series of the real symptom onset and the notification date, as reported by the ECDC, and that such delay is not constant in time.

## Results

For each time-series, we calculate the Time-Varying Reproduction Number R(t) by means of *EpiEstim*, a state-of-the-art method for the estimation of R(t), currently adopted by several national public health authorities [8]. Note that since surveillance data consolidates as time goes by, our R(t) estimations do not take into account the last month of data.

Figure 2 shows a comparison of the values obtained for R(t) for each of the time series considered. The comparison reveals remarkable differences between the different R(t) curves. Several observations, which hold for both countries, are in order. First, neither the incidence curve by notification date, nor the ones by inferred symptoms’ onset allow for a correct assessment of the early behavior of the epidemic. The curve obtained using the symptom onset inferred by convolution underestimates the true R(t) value, while those corresponding to notification dates and time-shifted dates of death greatly overestimate it (R(t) could be up to 6). Second, both of the inferred symptoms’ onset time-series yield R(t) curves which, past the dates of implementation of the containment measures, decline slower than the one obtained for the real symptoms’ onset. Third, only when using the time-series of real symptoms’ onset, R(t) falls below 1 within 5-6 days after national lockdowns in both countries, which is compatible with the incubation period of the disease [9]. On the contrary, by using notification dates, R(t) does not fall below 1 until early April. These dates are explicitly shown in Table 1.

**Table 1.** First day for which the value of R(t) is consistently below 1 within a 95% credible interval for each time-series. The results hold for sliding windows from 1 to 10 days.

|  | R(t)<1 |  |  |  | National lockdown |
| --- | --- | --- | --- | --- | --- |
|  | Notification | Inferred symptom onset A | Inferred symptom onset B | Real symptom onset |  |
| **Spain** | 31/03 | 25/03 | 25/03 | 21/03 | 15th of March |
| **Italy** | 28/03 | 22/03 | 21/03 | 17/03 | 12th of March |

**Figure 2.**
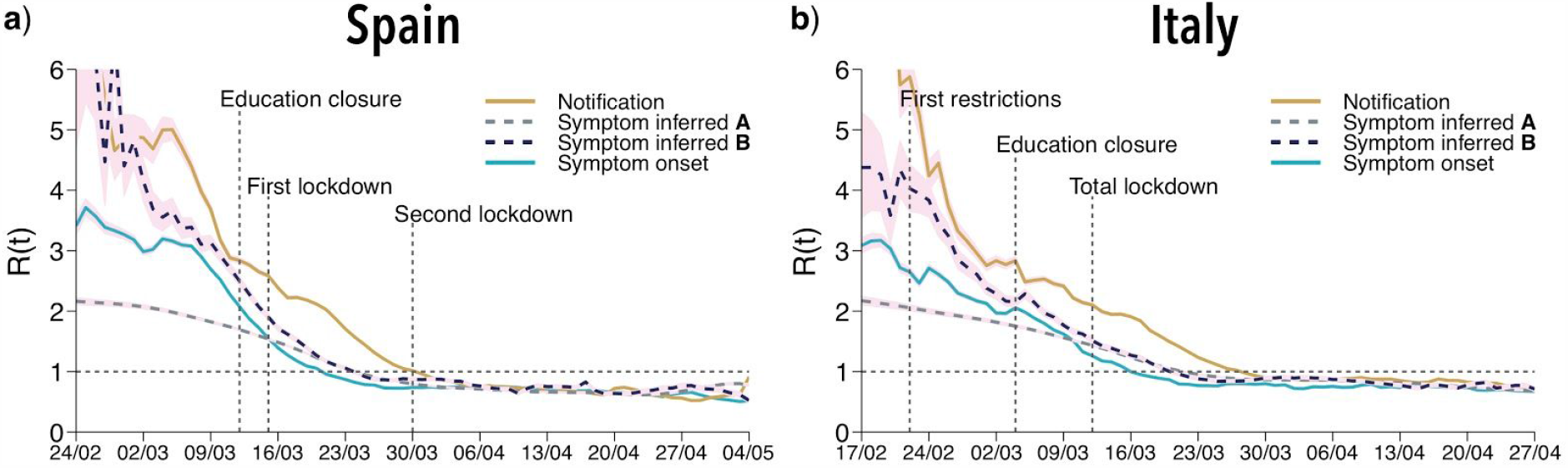
Estimated R(t) curves for Spain (left) and Italy (right) for each of the four time-series considered. We use EpiEstim with 7 days sliding windows and a gamma distributed serial interval with shape 1.87 and rate 0.28 [10]. 95% credible intervals are plotted as pink areas. The dashed vertical lines represent the dates in which main containment measures were applied in each country. For Spain, the “second lockdown” represents the all non-essential activity closure that was implemented on top of the lockdown imposed earlier.

## Conclusions

During the COVID-19 pandemic, in a much-needed effort to promote data availability, national health authorities of many countries have released, often on a daily basis and at a sub-national spatial granularity, the time-series of epidemiologically relevant quantities as measured through their surveillance systems. Such publicly-released data typically include the new number of laboratory-confirmed COVID-19 positives, hospitalized cases, and deaths. Although the dates of symptoms’ onset are also collected by surveillance systems for a fraction of cases, such information is often missing from publicly available data. For instance, until June 2020, Spanish authorities only released the aggregated time-series by symptoms’ onset in graphic format for the whole country. Italy, on the other hand, has periodically published this data for each region but until now only in graphic format. Finally, the ECDC currently only reports the values by date of notification.

Our study quantifies the biases in the estimation of R(t) introduced by using surveillance data different from the real dates of symptoms onset. In particular, we showed that inferring the symptoms’ onset time-series from the notification date does not capture the early dynamics of the epidemic. While it is known that the estimation of R(t) is sensitive to various input parameters, such as the distribution of the serial interval and the generation time, our study highlights the importance of relying on high-quality incidence data, rather than inferred epidemic time-series.

Given the past, present, and future importance of a precise estimation of the time-varying reproductive number for the management of the COVID-19 pandemic, our work calls for an increased transparency in the data that are released publicly by health authorities, which should make their best effort to collect and share incidence case counts by date of symptoms onset - even for a fraction of the total. The effects of critical policy decisions, including severe mobility restrictions and other containment measures, can indeed be correctly evaluated only by using accurate case-based surveillance data.

## Data Availability

All data used is publicly available

## Footnotes

1 Official report from the Spanish Ministry of Health of the current situation as of 20th of May of 2020 (in Spanish): https://www.isciii.es/QueHacemos/Servicios/VigilanciaSaludPublicaRENAVE/EnfermedadesTransmisibles/Documents/INFORMES/Informes%20COVID-19/Informe%20n%C2%BA%2033.%20An%C3%A1lisis%20de%20los%20casos%20de%20COVID-19%20hasta%20el%2010%20de%20mayo%20en%20Espa%C3%B1a%20a%2029%20de%20mayo%20de%202020.pdf

2 Official report from the Italian Ministry of Health of the current situation as of 11th of June of 2020 (in Italian) https://www.epicentro.iss.it/coronavirus/bollettino/Report-COVID-2019_11_giugno.pdf

